## Supplementary Table 1; Supplementary Table 2; Supplementary Table 3; Supplementary Table 4; Supplementary Table 5; Supplementary Table 6 for "Increased Burden of Familial-associated Early-onset Cancer Risk among Minority Americans Compared to non-Latino Whites"

**Supplemental materials**

**Supplementary Table 1. Relative risks of the same type of early-onset cancer with the proband among siblings and mothers by ethnic group, 1989 to 2015, California, USA.**

| | Overall | | | Non-Latino White | | | Latino all races | | | |
|---|---|---|---|---|---|---|---|---|---|---|
| Cancer of the proband [†] | No. of Probands | No. of affected relatives [§] | SIR (95% CI) | No. of Probands | No. of relatives [§] | SIR (95% CI) | No. of Probands | No. of affected relatives [§] | SIR (95% CI) | p-value [‡] |
| Hematologic cancers[¶] | 11404 | 22 | 2.68* (1.68, 4.06) | 2702 | 6 | 2.64 (0.97, 5.76) | 5484 | 7 | 1.56 (0.63, 3.21) | 0.505 |
| Solid cancers[¶] | 17849 | 112 | 6.78* (5.58, 8.16) | 5471 | 31 | 4.41* (2.99, 6.25) | 7323 | 50 | 7.94* (5.89, 10.47) | 0.012 |
| Leukemias | 8500 | 11 | 2.48* (1.24, 4.45) | 1646 | ≦5 | 2.19 (0.27, 7.91) | 4374 | ≦5 | 1.45 (0.4, 3.72) | 0.996 |
| Lymphomas | 2928 | ≦5 | 5.9* (1.61, 15.11) | 1060 | ≦5 | 9.04* (1.86, 26.41) | 1121 | 0 | | NA |
| CNS tumors | 5739 | 11 | 6.19* (3.09, 11.08) | 1728 | ≦5 | 5.07* (1.38, 12.98) | 2242 | ≦5 | 4.66 (0.96, 13.61) | 0.788 |
| Neuroblastoma | 1445 | ≦5 | 10.31 (0.26, 57.44) | 367 | 0 | NA | 490 | 0 | | NA |
| Retinoblastoma | 718 | 10 | 454.55* (217.97, 835.93) | 126 | ≦5 | 200* (5.06, 1114.32) | 327 | 7 | 636.36* (255.85, 1311.15) | 0.446 |
| Renal tumors | 1069 | ≦5 | 15.87 (0.4, 88.44) | 209 | 0 | NA | 442 | ≦5 | 38.46 (0.97, 214.29) | NA |
| Hepatic tumors | 414 | 0 | NA | 74 | 0 | NA | 200 | 0 | | NA |
| Malignant bone tumors | 249 | 0 | NA | 73 | 0 | NA | 121 | 0 | | NA |
| Sarcomas | 2915 | 10 | 31.45* (15.08, 57.83) | 816 | ≦5 | 18.87* (2.28, 68.16) | 1259 | ≦5 | 35.97* (11.68, 83.94) | 0.687 |
| GCT | 2423 | ≦5 | 7.71* (1.59, 22.54) | 672 | 0 | NA | 1270 | ≦5 | 8.62* (1.04, 31.14) | NA |
| Epithelial neoplasms | 3050 | 24 | 40.68* (26.06, 60.53) | 1458 | 9 | 21.33* (9.75, 40.49) | 1046 | 9 | 55.56* (25.4, 105.46) | 0.065 |
| Other | 255 | 0 | NA | 79 | 0 | NA | 90 | 0 | NA | NA |

[†] Cancers were classified into subgroups as defined by the International Classification of Childhood Cancer, Third edition (ICCC-3, November 2012) (https://seer.cancer.gov/iccc/iccc3.html).
[§] Affected relatives include mother and siblings of the proband diagnosed with early-onset cancer under 26 years of age.
[‡] p-value comparing the SIRs between non-Latino White and Latino all races using an approximate Chi-squared test.
[¶] Hematologic cancers include group 1, leukemias, myeloproliferative diseases, and myelodysplastic diseases; group 2, lymphomas and reticuloendothelial neoplasms. Solid cancers include I group 3, CNS and miscellaneous intracranial and intraspinal neoplasms; group 4, neuroblastoma and other peripheral nervous cell tumors; group 5, retinoblastoma; group 6, renal tumors; group 7, hepatic tumors; group 8, malignant bone tumors; group 9, soft tissue and other extraosseous sarcomas; group 10, germ cell tumors, trophoblastic tumors, and neoplasms of gonads; group 11, other malignant epithelial neoplasms and malignant melanomas; group 12, other and unspecified malignant neoplasms.
SIR, Standardized incidence ratio. CI, confidence interval.
* Statistically significant standardized incidence ratio with $p<0.05$ assuming a Poisson distribution.

**Supplemental Table 2. Relative risks of siblings and mothers for a specific type of early-onset cancer (diagnosed 0 to 26 years of age) given a proband with cancer, 1989 to 2015, California, USA.**

| Cancer of the proband † | Cancer of the relative § | No. of affected relatives § | SIR (95% CI) |
|---|---|---|---|
| | Leukemias | 11 | 2.48* (1.24, 4.45) |
| | Lymphomas | 6 | 2.46 (0.9, 5.35) |
| | CNS tumors | 6 | 2.02 (0.74, 4.4) |
| | Hepatic tumors | ≦5 | 0.41 (0.01, 2.28) |
| | Sarcomas | 8 | 7.36* (3.18, 14.5) |
| | GCT | 6 | 3.51* (1.29, 7.64) |
| **Leukemias** | Epithelial neoplasms | 13 | 5.37* (2.86, 9.18) |
| | Leukemias | 7 | 7.47* (3, 15.39) |
| | Lymphomas | ≦5 | 5.9* (1.61, 15.11) |
| | CNS tumors | ≦5 | 2.89 (0.35, 10.46) |
| | Neuroblastoma | ≦5 | 15.38 (0.39, 85.72) |
| | Retinoblastoma | ≦5 | 40* (1.01, 222.86) |
| | Renal tumors | ≦5 | 12.99 (0.33, 72.36) |
| | Sarcomas | ≦5 | 7.07 (0.86, 25.53) |
| | GCT | ≦5 | 1.93 (0.05, 10.78) |
| **Lymphomas** | Epithelial neoplasms | ≦5 | 2.67 (0.32, 9.65) |
| | Leukemias | ≦5 | 1.94 (0.63, 4.53) |
| | Lymphomas | ≦5 | 1.31 (0.16, 4.72) |
| | CNS tumors | 11 | 6.19* (3.09, 11.08) |
| | Renal tumors | ≦5 | 4.29 (0.11, 23.91) |
| | Sarcomas | 6 | 8.97* (3.29, 19.52) |
| | GCT | ≦5 | 1.81 (0.22, 6.55) |
| | Epithelial neoplasms | 10 | 6.37* (3.05, 11.71) |
| **CNS tumors** | Other | ≦5 | 0.65 (0.02, 3.64) |

| | | | |
|---|---|---|---|
| | Lymphomas | ≦5 | 2.23 (0.06, 12.41) |
| | Neuroblastoma | ≦5 | 10.31 (0.26, 57.44) |
| | Sarcomas | ≦5 | 4.78 (0.12, 26.66) |
| | GCT | ≦5 | 3.33 (0.08, 18.57) |
| | Epithelial neoplasms | ≦5 | 5.09 (0.62, 18.38) |
| **Neuroblastoma** | Other | ≦5 | 2.23 (0.06, 12.41) |
| | Lymphomas | ≦5 | 4.5 (0.11, 25.1) |
| | Retinoblastoma | 10 | 454.55* (217.97, 835.93) |
| | Malignant bone tumors | ≦5 | 4.5 (0.11, 25.1) |
| | Sarcomas | ≦5 | 9.52 (0.24, 53.06) |
| **Retinoblastoma** | Epithelial neoplasms | ≦5 | 5.24 (0.13, 29.17) |
| | Lymphomas | ≦5 | 3.02 (0.08, 16.83) |
| | CNS tumors | ≦5 | 2.37 (0.06, 13.2) |
| | Renal tumors | ≦5 | 15.87 (0.4, 88.44) |
| | Sarcomas | ≦5 | 13.33* (1.61, 48.16) |
| | GCT | ≦5 | 4.42 (0.11, 24.65) |
| **Renal tumors** | Epithelial neoplasms | ≦5 | 9.17* (1.89, 26.81) |
| | Leukemias | ≦5 | 4.08 (0.1, 22.74) |
| **Hepatic tumors** | Epithelial neoplasms | ≦5 | 9.52 (0.24, 53.06) |
| | Leukemias | ≦5 | 14.49 (0.37, 80.75) |
| **Malignant bone tumors** | Epithelial neoplasms | ≦5 | 16.39 (0.41, 91.34) |
| | Leukemias | 8 | 7.01* (3.03, 13.82) |
| | Lymphomas | ≦5 | 1.37 (0.03, 7.61) |
| | CNS tumors | 7 | 8.62* (3.47, 17.76) |
| | Neuroblastoma | ≦5 | 11.11 (0.28, 61.91) |
| | Renal tumors | ≦5 | 19.8* (2.4, 71.53) |
| | Sarcomas | 10 | 31.45* (15.08, 57.83) |
| | GCT | ≦5 | 7.41* (2.02, 18.97) |
| **Sarcomas** | Epithelial neoplasms | 6 | 7.56* (2.77, 16.45) |

| | | | |
|---|---|---|---|
| | Other | ≦5 | 1.37 (0.03, 7.61) |
| | Leukemias | ≦5 | 5.81* (1.58, 14.89) |
| | Lymphomas | ≦5 | 4.06 (0.49, 14.65) |
| | CNS tumors | ≦5 | 5.93* (1.22, 17.33) |
| | Neuroblastoma | ≦5 | 19.23 (0.49, 107.15) |
| | Renal tumors | ≦5 | 16.67 (0.42, 92.86) |
| | Sarcomas | ≦5 | 9.66* (1.17, 34.9) |
| | GCT | ≦5 | 7.71* (1.59, 22.54) |
| **GCT** | Epithelial neoplasms | ≦5 | 6.92* (1.89, 17.72) |
| | Leukemias | ≦5 | 8.39* (2.28, 21.47) |
| | Lymphomas | ≦5 | 2.19 (0.06, 12.22) |
| | CNS tumors | ≦5 | 7.56* (1.56, 22.08) |
| | Neuroblastoma | ≦5 | 41.67* (1.05, 232.15) |
| | Sarcomas | ≦5 | 10.99* (1.33, 39.7) |
| | GCT | ≦5 | 2.63 (0.07, 14.66) |
| **Epithelial neoplasms** | Epithelial neoplasms | 24 | 40.68* (26.06, 60.53) |
| | Lymphomas | ≦5 | 16.39 (0.41, 91.34) |
| | CNS tumors | ≦5 | 14.29 (0.36, 79.59) |
| | Neuroblastoma | ≦5 | 125* (3.16, 696.45) |
| **Other** | Sarcomas | ≦5 | 37.04 (0.94, 206.36) |

[†] Cancers were classified into subgroups as defined by the International Classification of Childhood Cancer, Third edition (ICCC-3, November 2012) (https://seer.cancer.gov/iccc/iccc3.html).
[§] Affected relatives include mother and siblings of the proband diagnosed with early-onset cancer under 26 years of age.
SIR, Standardized incidence ratio. CI, confidence interval.
* Statistically significant standardized incidence ratio with $p<0.05$ assuming a Poisson distribution.

**Supplemental Table 3. Relative risks of any early-onset cancer (diagnosed at 0 to 26 years of age) for siblings and mothers of the same type of cancer with the proband given a proband with cancer by subgroups, 1989 to 2015, California, USA.**

| | | All cancers | | The same type of cancer | |
|---|---|---|---|---|---|
| **Cancer of the proband** † | **No. of probands** | **No. of affected relatives** § | **SIR (95% CI)** | **No. of affected relatives** § | **SIR (95% CI)** |
| **Lymphoid leukemias** | 6705 | 25 | 2.7* (1.91, 3.7) | ≦5 | 1.86 (0.6, 4.34) |
| **Acute myeloid leukemias** | 1317 | 9 | 4.61* (2.46, 7.89) | ≦5 | 27.27* (5.62, 79.7) |
| **Hodgkin lymphomas** | 1399 | 9 | 7.12* (3.79, 12.17) | ≦5 | 10.53* (1.27, 38.02) |
| **Non-Hodgkin lymphomas (except Burkitt lymphoma)** | 985 | ≦5 | 4.02* (1.48, 8.75) | ≦5 | 11.11 (0.28, 61.91) |
| **Ependymomas and choroid plexus tumor** | 561 | ≦5 | 2.46 (0.51, 7.19) | 0 | NA |
| **Astrocytomas** | 1965 | 11 | 3.6* (1.97, 6.03) | ≦5 | 7.69 (0.93, 27.79) |
| **Intracranial and intraspinal embryonal tumors** | 1010 | 6 | 2.67* (1.07, 5.5) | ≦5 | 11.11 (0.28, 61.91) |
| **Other gliomas** | 815 | 8 | 5.34* (2.31, 10.53) | ≦5 | 66.67* (8.07, 240.82) |
| **Other specified intracranial and intraspinal neoplasms** | 1470 | ≦5 | 5.48* (2.5, 10.4) | ≦5 | 22.22* (2.69, 80.27) |
| **Neuroblastoma and ganglioneuroblastoma** | 1409 | 6 | 2.06 (0.83, 4.25) | ≦5 | 9.09 (0.23, 50.65) |
| **Nephroblastoma and other nonepithelial renal tumors** | 1032 | 6 | 3.67* (1.68, 6.97) | ≦5 | 16.67 (0.42, 92.86) |
| **Rhabdomyosarcomas** | 719 | 11 | 8.46* (4.63, 14.2) | 0 | NA |
| **Fibrosarcomas to peripheral nerve sheath tumors to and other fibrous neoplasms** | 434 | 7 | 10.46* (4.21, 21.56) | ≦5 | 200* (24.22, 722.47) |
| **Other specified soft tissue sarcomas** | 1624 | 11 | 7.57* (4.56, 11.82) | ≦5 | 55.56* (18.04, 129.65) |
| **Malignant gonadal germ cell tumors** | 1030 | 12 | 5.44* (3.05, 8.97) | ≦5 | 6.67 (0.81, 24.08) |
| **Malignant melanomas** | 788 | ≦5 | 3.65 (0.75, 10.68) | ≦5 | 25 (0.63, 139.29) |

| | | | | | |
|---|---|---|---|---|---|
| **Other and unspecified carcinomas** | 1845 | 20 | 18.11* (12.13, 26.01) | 17 | 113.33* (66.02, 181.46) |

†Cancers were classified into subgroups as defined by the International Classification of Childhood Cancer, Third edition (ICCC-3, November 2012) (https://seer.cancer.gov/iccc/iccc3.html).

§ Affected relatives include mother and siblings of the proband diagnosed with early-onset cancer under 26 years of age.

SIR, Standardized incidence ratio. CI, confidence interval.

* Statistically significant standardized incidence ratio with $p<0.05$ assuming a Poisson distribution.

**Supplementary Table 4. Relative risks of any early-onset cancer (diagnosed 0 to 26 years of age) among siblings and mothers by ethnic group, 1989 to 2015, California, USA.**

| | Non-Latino White | | | Latino all races | | | | Non-Latino API | | | | Non-Latino Black | | | |
|---|---|---|---|---|---|---|---|---|---|---|---|---|---|---|---|
| Cancer of the proband † | No. of Probands | No. of affected relatives § | SIR (95% CI) | No. of Probands | No. of affected relatives § | SIR (95% CI) | p-value ‡ | No. of Probands | No. of affected relatives § | SIR (95% CI) | p-value ‡ | No. of Probands | No. of affected relatives § | SIR (95% CI) | p-value ‡ |
| Overall | 8119 | 50 | 2.6* (1.93, 3.43) | 12736 | 78 | 3.36* ( 2.66, 4.19) | 0.183 | 1677 | 11 | 4.58* (2.29, 8.2) | 0.128 | 1102 | 13 | 6.96* (3.71, 11.91) | 0.002 |
| Hematologic cancers¶ | 2702 | 19 | 2.69* (1.62, 4.2) | 5484 | 27 | 2.48* (1.64, 3.61) | 0.910 | 679 | 8 | 7.56* (3.26, 14.9) | 0.023 | 391 | ≦5 | 6.14* (1.67, 15.73) | 0.242 |
| Solid cancers¶ | 5471 | 37 | 3.02* (2.12, 4.16) | 7323 | 62 | 4.98* (3.82, 6.39) | 0.019 | 1021 | 7 | 5.07* (2.04, 10.44) | 0.306 | 719 | 9 | 7.35* (3.36, 13.95) | 0.026 |
| Leukemias | 1646 | 11 | 2.11* (1.05, 3.77) | 4374 | 21 | 2.35* (1.45, 3.59) | 0.911 | 457 | 6 | 6.94* (2.55, 15.12) | 0.032 | 205 | ≦5 | 6.58* (1.36, 19.22) | 0.176 |
| Lymphomas | 1060 | 8 | 4.32* (1.87, 8.52) | 1121 | 8 | 4.09* (1.77, 8.07) | 0.887 | 226 | ≦5 | 20.3* (5.53, 51.98) | 0.022 | 187 | ≦5 | 5.13 (0.13, 28.53) | 0.685 |
| CNS tumors | 1728 | 12 | 2.63* (1.36, 4.59) | 2242 | 12 | 2.86* (1.48, 4.99) | 0.999 | 311 | ≦5 | 7.14* (1.47, 20.87) | 0.250 | 239 | ≦5 | 7.56* (1.56, 22.08) | 0.216 |
| Neuroblastoma | 367 | ≦5 | 0.7 (0.02, 3.92) | 490 | ≦5 | 1.7 (0.21, 6.13) | 0.871 | 51 | ≦5 | 6.54 (0.17, 36.37) | 0.466 | 48 | 0 | NA | NA |
| Retinoblastoma | 126 | ≦5 | 3.65 (0.44, 13.18) | 327 | 9 | 12.31* (5.63, 23.37) | 0.178 | 35 | 0 | NA | NA | 30 | ≦5 | 11.49 (0.29, 63.95) | 0.881 |
| Renal tumors | 209 | ≦5 | 2.64 (0.32, 9.54) | 442 | ≦5 | 2.91 (0.6, 8.5) | 0.728 | 28 | 0 | NA | NA | 53 | ≦5 | 8.85 (0.22, 49.24) | 0.850 |
| Hepatic tumors | 74 | 0 | NA | 200 | ≦5 | 2.43 (0.06, 13.52) | NA | 13 | 0 | NA | NA | 6 | 0 | NA | NA |
| Bone tumors | 73 | 0 | NA | 121 | ≦5 | 6.06 (0.15, 33.77) | NA | 13 | 0 | NA | NA | 17 | 0 | NA | NA |

| | | | | | | | | | | | | | | | |
|---|---|---|---|---|---|---|---|---|---|---|---|---|---|---|---|
| Sarcomas | 816 | 7 | 3.77* (1.51, 7.76) | 1259 | 20 | 9.05* (5.53, 13.98) | 0.062 | 160 | ≦5 | 4.46 (0.11, 24.84) | 0.681 | 132 | ≦5 | 10.68* (2.2, 31.19) | 0.267 |
| GCT | 672 | ≦5 | 3.42 (0.93, 8.77) | 1270 | 10 | 6.2* (2.97, 11.41) | 0.454 | 184 | ≦5 | 4.85 (0.12, 27.01) | 0.755 | 70 | 0 | NA | NA |
| Epithelial neoplasms | 1458 | 14 | 8.6* (4.7, 14.43) | 1046 | 15 | 15.15* (8.48, 24.99) | 0.176 | 239 | ≦5 | 13.25* (1.6, 47.82) | 0.899 | 122 | ≦5 | 22.47* (2.72, 81.13) | 0.450 |
| Other | 79 | ≦5 | 10.47* (1.27, 37.83) | 90 | ≦5 | 5.85 (0.15, 32.58) | 0.924 | 8 | 0 | NA | NA | 19 | 0 | NA | NA |

[†] Cancers were classified into subgroups as defined by the International Classification of Childhood Cancer, Third edition (ICCC-3, November 2012) (https://seer.cancer.gov/iccc/iccc3.html).
[§] Affected relatives include mother and siblings of the proband diagnosed with early-onset cancer under 26 years of age.
[‡] p-value comparing the SIRs between non-Latino White and Latino all races using an approximate Chi-squared test.
[¶] Hematologic cancers include group 1, leukemias, myeloproliferative diseases, and myelodysplastic diseases; group 2, lymphomas and reticuloendothelial neoplasms. Solid cancers include I group 3, CNS and miscellaneous intracranial and intraspinal neoplasms; group 4, neuroblastoma and other peripheral nervous cell tumors; group 5, retinoblastoma; group 6, renal tumors; group 7, hepatic tumors; group 8, malignant bone tumors; group 9, soft tissue and other extraosseous sarcomas; group 10, germ cell tumors, trophoblastic tumors, and neoplasms of gonads; group 11, other malignant epithelial neoplasms and malignant melanomas; group 12, other and unspecified malignant neoplasms.
SIR, Standardized incidence ratio. CI, confidence interval.
* Statistically significant standardized incidence ratio with $p<0.05$ assuming a Poisson distribution.

**Supplemental Table 5. Second primary malignancies in families exhibited familial risks and families did not exhibit familial risk.**

| | Family exhibited familial risks | Family did not exhibit familial risk |
|---|---|---|
| **No. of SPMs** | 14 | 373 |
| **Total No. of family members** | 2,432 | 119,136 |
| **No. of SPMs per family member** † | 0.58% | 0.31% |
| **Average family size** | 4.35 people/family | 4.17 people/family |
| **Average time between FPM and SPM diagnosis** | 6.69 years | 6.35 years |
| **Average time from FPM diagnosis to the end of study (2015)** | 12.23 years | 11.74 years |

FPM, first primary malignancy. SPM, second primary malignancy.
† No. of SPMs per family member was compared with a Chi-squared test. Families that exhibited familial risks have higher proportion of SPMs compared to families that did not exhibit familial risk ($Chi^2$= 5.13; p= 0.023).

**Supplemental Table 6. Relative risks of second primary malignancies of the same type of early-onset cancer (diagnosed 0 to 26 years of age) with the first primary malignancy by ethnic groups, California, USA.**

| | Overall | | | Non-Latino White | | | Latino (all races) | | | |
|---|---|---|---|---|---|---|---|---|---|---|
| **Cancer of the proband** † | **No. of FPMs** | **No. of SPMs** | **SIR (95% CI)** | **No. of FPMs** | **No. of SPMs** | **SIR (95% CI)** | **No. of FPMs** | **No. of SPMs** | **SIR (95% CI)** | **p-value** ‡ |
| **Hematologic cancers¶** | 11,553 | 50 | 6.1* (4.53, 8.04) | 2,727 | 15 | 6.61* (3.7, 10.9) | 5,528 | 19 | 4.23* (2.55, 6.61) | 0.262 |
| **Solid cancers¶** | 18,893 | 30 | 1.82* (1.22, 2.59) | 5,673 | 13 | 1.85 (0.98, 3.16) | 7,551 | 13 | 2.06* (1.1, 3.53) | 0.938 |
| **Leukemias** | 8,543 | 41 | 9.26* (6.65, 12.56) | 1,649 | 11 | 12.05* (6.01, 21.56) | 4,385 | 16 | 5.82* (3.32, 9.44) | 0.094 |
| **Lymphomas** | 2,960 | 9 | 13.27* (6.07, 25.2) | 1,063 | ≦5 | 12.05* (3.28, 30.85) | 1,124 | ≦5 | NA | 0.768 |
| **CNS tumors** | 5,877 | 29 | 16.32* (10.93, 23.44) | 1,765 | 12 | 15.21* (7.86, 26.57) | 2,277 | 11 | 17.08* (8.53, 30.56) | 0.943 |
| **Neuroblastoma** | 1,450 | ≦5 | 20.62* (2.5, 74.48) | 369 | ≦5 | NA | 491 | 0 | NA | NA |
| **Retinoblastoma** | 728 | 0 | NA | 127 | 0 | NA | 334 | 0 | NA | NA |
| **Renal tumors** | 1,078 | 0 | NA | 209 | 0 | NA | 444 | 0 | NA | NA |
| **Hepatic tumors** | 417 | 0 | NA | 74 | 0 | NA | 200 | 0 | NA | NA |
| **Malignant bone tumors** | 250 | 0 | NA | 73 | 0 | NA | 121 | 0 | NA | NA |
| **Sarcomas** | 3,028 | 16 | 50.31* (28.76, 81.71) | 850 | ≦5 | 47.17* (15.32, 110.08) | 1,297 | 7 | 50.36* (20.25, 103.76) | 0.858 |
| **GCT** | 2,468 | ≦5 | 5.14 (0.62, 18.57) | 679 | 0 | NA | 1,287 | ≦5 | 8.62* (1.04, 31.14) | NA |
| **Epithelial neoplasms** | 3,479 | 10 | 16.95* (8.13, 31.17) | 1,489 | 6 | 14.22* (5.22, 30.95) | 1,057 | ≦5 | 24.69* (6.73, 63.22) | 0.605 |
| **Other** | 261 | 0 | NA | 79 | 0 | NA | 90 | 0 | NA | NA |

SIR, Standardized incidence ratio. CI, confidence interval. FPM, first primary malignancy. SPM, second primary malignancy.
† Cancers were classified into subgroups as defined by the International Classification of Childhood Cancer, Third edition (ICCC-3, November 2012) (https://seer.cancer.gov/iccc/iccc3.html).
‡ p-value comparing the SIRs between non-Latino White and Latino all races using an approximate Chi-squared test.
¶ Hematologic cancers include group 1, leukemias, myeloproliferative diseases, and myelodysplastic diseases; group 2, lymphomas and reticuloendothelial neoplasms. Solid cancers include I group 3, CNS and miscellaneous intracranial and intraspinal neoplasms; group 4, neuroblastoma and other peripheral nervous cell tumors; group 5, retinoblastoma; group 6, renal tumors; group 7, hepatic tumors; group 8,

malignant bone tumors; group 9, soft tissue and other extraosseous sarcomas; group 10, germ cell tumors, trophoblastic tumors, and neoplasms of gonads; group 11, other malignant epithelial neoplasms and malignant melanomas; group 12, other and unspecified malignant neoplasms.
SIR, Standardized incidence ratio. CI, confidence interval. FPM, first primary malignancy. SPM, second primary malignancy.
* Statistically significant standardized incidence ratio with $p<0.05$ assuming a Poisson distribution.
