## Supplementary material for "Increased Burden of Familial-associated Early-onset Cancer Risk among Minority Americans Compared to non-Latino Whites": Source Code File

### Statistics and coding supplement: Familial_risk_CCR_eLife

Qianxi Feng

12/7/2020

### 0. Pre-processing

#### 0.1 Load required packages and read file

```
rm(list = ls())
library(descr)
library(dplyr)
```

```
##
## Attaching package: 'dplyr'
```

```
## The following objects are masked from 'package:stats':
##
##     filter, lag
```

```
## The following objects are masked from 'package:base':
##
##     intersect, setdiff, setequal, union
```

```
library(tidyverse)
```

```
## ── Attaching packages ─────────────────────────────────────── tidyverse 1.3.0 ──
```

```
## ✓ ggplot2 3.3.3     ✓ purrr   0.3.4
## ✓ tibble  3.1.0     ✓ stringr 1.4.0
## ✓ tidyr   1.1.3     ✓ forcats 0.5.1
## ✓ readr   1.4.0
```

```
## ── Conflicts ────────────────────────────────────────── tidyverse_conflicts() ──
## x dplyr::filter() masks stats::filter()
## x dplyr::lag()    masks stats::lag()
```

```
library(data.table)
```

```
##
## Attaching package: 'data.table'
```

```
## The following object is masked from 'package:purrr':
##
##     transpose
```

```
## The following objects are masked from 'package:dplyr':
##
##     between, first, last
```

```
library(readxl)
workdir<- "/Volumes/projects_wiemels/V_Foundation/SenkeiF/Codes&Intermediate datasets/"
fambirthccrT<- read.csv(paste0(workdir, "Family_CCR_birth.csv"))
```

### 0.2 Recode variables

```
# Create a numeric child id variable, a numeric mother_id variable, and a numeric patien
t_id variable
fambirthccrT$vitstat_childID_num<- as.numeric(as.character(gsub( "_", "", fambirthccrT$v
itstat_childID)))
# summary(fambirthccrT$vitstat_childID_num)
fambirthccrT$mother_id_num<- as.numeric(as.character(fambirthccrT$mother_id))
# summary(fambirthccrT$mother_id_num)
fambirthccrT$PATIENT_ID_num<- as.numeric(as.character(fambirthccrT$PATIENT_ID))

# change YEARDX and birth_year to numeric
fambirthccrT[c("YEARDX", "birth_year")] <- lapply(fambirthccrT[c("YEARDX", "birth_year"
)], as.character)
fambirthccrT[c("YEARDX", "birth_year")] <- lapply(fambirthccrT[c("YEARDX", "birth_year"
)], as.numeric)
class(fambirthccrT$YEARDX)
```

```
## [1] "numeric"
```

```
class(fambirthccrT$birth_year)
```

```
## [1] "numeric"
```

### 0.3 Create functions and data subsets for future use

```
# Keep complete rows based on certain columns
completeFun <- function(data, desiredCols) {
  completeVec <- complete.cases(data[, desiredCols])
  return(data[completeVec, ])
}

# Data set of cases
fambirthccrTcases<- fambirthccrT[!is.na(fambirthccrT$PATIENT_ID_num), ]

# Data set of child cases
childCasesdf<- completeFun(fambirthccrTcases, c("vitstat_childID_num")) #30,019 child ca
ses

# Data set of adult cases
adultCasesdf<- fambirthccrTcases[is.na(fambirthccrTcases$vitstat_childID_num), ]
motherCasesdf<- subset(adultCasesdf, status == "Mother") # 453 patients
motherCasesdf<- subset(motherCasesdf, AGE>=14) # mothers > age 14
```

### 0.4 Prepare the ICD codes

Coding criteria: https://seer.cancer.gov/iccc/iccc3.html (https://seer.cancer.gov/iccc/iccc3.html)

```
icd<- read.csv(paste0(workdir, "icd codes 12 broad groups.csv"))
# ICD codes by group
icd_list <- list()
for (i in 1:12){
 icd_list[[i]] <- c(icd$icd_codes[icd$sitegroup == i])
}

# ICD codes for hematological cancers and solid cancers
hemaICD<- c(icd$icd_codes[icd$sitegroup<=2])
solidICD<- c(icd$icd_codes[icd$sitegroup>=3])
```

### 1. Standardized incidence ratio (SIR) calculation for affected mother/sibling of any cancer

The calculation of SIR follows following the equation:

$$SIR = \frac{O}{E} = \frac{\sum_{i=1}^{N} \sum_{j=1}^{n_i} D_{ij}}{\sum_{i=1}^{N} \sum_{j=1}^{n_i} \sum_{k=1}^{K_{max}} \lambda_k t_{ijk}}$$

Where N is the number of families, $n_i$ is the number of non-proband individuals of interest (siblings/SPMs/mothers) in family i, and $K_{max}$ is the total number of age intervals. The data for each individual includes a disease indicator ($D_{ij}$) and the number of years “at risk” during the kth age interval ($t_{ijk}$). A given individual is defined to be at risk beginning at their age when the proband in their family is diagnosed and ending either when they become affected themselves or they are censored due to end of study follow-up. For siblings and mothers, age was stratified into seven groups as 0, 0-4, 5-9, 10-14, 15-19, 20-24 and 25-29 years. For the calculation of SIRs within a given race group, $\lambda_k$ is the race-, sex- and age-specific incidence rate of a given cancer.

### 1.1 SIR overall

- Count the observed number of cases
- Use healthy siblings and mothers as the denominator

1. Year at risk of a sibling =

- 2015- year of proband's diagnosis if sibling was born before proband was diagnosed;
- 2015- birth_year of sibling if sibling was born after proband was diagnosed

2. Year at risk of a mother =

- 2015- year of proband's diagnosis

```
## Any cancer
any_cancer <- function(icd, race, ethnicity){
  cases<- subset(fambirthccrTcases, HISTO_T3 %in% icd & RaceEthnicityOfMother %in% race
 & HispanicOriginCodeOfMother %in% ethnicity)

  # Restrict yeardx to 1989-2015
  cases<- subset(cases, YEARDX>=1989& YEARDX<=2015)
  casesN<- nrow(cases) # Count number of cases

  # Find the probands
  ## Probands are any kid who got the case, one family can only have one proband (case o
f the earliest yeardx)
  probands_pre<- completeFun(cases, c("vitstat_childID_num")) # Extract child cases
  probands_pre<- probands_pre[order(probands_pre$YEARDX), ] # Sort by yeardx
  probands<- distinct(probands_pre, mother_id_num, .keep_all= TRUE) # Keep only one chil
d for each family
  fams<- subset(fambirthccrT, mother_id_num %in% c(probands$mother_id_num))
  probandsN<- nrow(data.frame(table(probands$vitstat_childID_num))) # count number of pr
obands

#############################################
  # Count the number of mother cases of any cancer, diagnosed under 26
  mothers<- subset(fams, status == "Mother")
  motherCasesdf<- completeFun(mothers, c("PATIENT_ID_num"))
  # motherless14<- subset(motherCasesdf, AGE<14)
  motherCasesdf<- subset(motherCasesdf, AGE<=26)
  motherCases_num_rec<- nrow(motherCasesdf)
  motherCases_num_people<- nrow(data.frame(table(motherCasesdf$PATIENT_ID_num)))
  ############ motherCases_num_people is part of the numerator ############

#############################################
  # Count the number of sibling cases of any cancer
  childCases<- completeFun(fams, c("vitstat_childID_num", "PATIENT_ID_num"))
  sibCasesdf<- subset(childCases, !(childCases$PATIENT_ID_num %in% c(probands$PATIENT_ID
_num)))
  sibCase_num_rec<- nrow(sibCasesdf)
  sibCase_num_people<- nrow(data.frame(table(sibCasesdf$PATIENT_ID_num)))
  ############ sibCase_num_people is part of the numerator ############

#############################################
#Find healthy siblings to calculate the denominator
  children<- completeFun(fams, c("vitstat_childID_num"))
  allSib<- subset(children, !(PATIENT_ID_num %in% c(probands$PATIENT_ID_num)))
  #all Siblings (including healthy)

# year at risk of a sibling=
## 1) 2015- year of proband's diagnosis if sibling was born before proband was diagnosed
## 2) 2015- birth_year of sibling if sibling was born after proband was diagnosed
  probands_1<- probands[order(probands$mother_id, probands$AGE), ]
  sum(unique(length(probands_1$mother_id_num)))
  probands_1<- probands_1[!duplicated(probands_1$mother_id_num),]
  probands_1$status_new<- "Proband"
  allSib$status_new<- "Sibling"
```

```
  children_1<- rbind(probands_1, allSib)
  children_1<- children_1[order(children_1$mother_id_num), ]

  setDT(children_1)[, risk_status := birth_year- YEARDX[status_new=="Proband"], by = mot
her_id_num]
  children_1$risk_statusN[children_1$risk_status<=0]<-1 #birth_year<=yeardx
  children_1$risk_statusN[children_1$risk_status>0]<-2 #birth_year>yeardx

  #1)
  children_11<- subset(children_1, risk_statusN==1)
  setDT(children_11)[, years_at_risk := 2015- YEARDX[status_new=="Proband"], by = mother
_id_num]
  setDT(children_11)[, AgeDiff := birth_year- birth_year[status_new=="Proband"], by = mo
ther_id_num]
  children_11Sib<- subset(children_11, status_new=="Sibling")
#2)
  children_12<- subset(children_1, risk_statusN==2)
  children_12$years_at_risk<- 2015-children_12$birth_year
  children_12$AgeDiff<- 0
  children_1<- rbind(children_11, children_12)

  # Crop out siblings
  allSib<- subset(children_1, status_new=="Sibling")
  allSib$age_sib<- 2015-allSib$birth_year

  # Recode age
  allSib$agegp[allSib$age_sib==0]<- "00 years"
  allSib$agegp[allSib$age_sib>=1&allSib$age_sib<=4]<- "01-04 years"
  allSib$agegp[allSib$age_sib>=5&allSib$age_sib<=9]<- "05-09 years"
  allSib$agegp[allSib$age_sib>=10&allSib$age_sib<=14]<- "10-14 years"
  allSib$agegp[allSib$age_sib>=15&allSib$age_sib<=19]<- "15-19 years"
  allSib$agegp[allSib$age_sib>=20&allSib$age_sib<=24]<- "20-24 years"
  allSib$agegp[allSib$age_sib>=25]<- "25-29 years"

  # Keep only healthy siblings to the denominator
  summary(allSib$PATIENT_ID)
  healthySib<- subset(allSib, !(vitstat_childID_num %in% c(sibCasesdf$vitstat_childID_nu
m)))
  healthySib$Count<- 1
  healthySib_aggr<- aggregate(Count~ agegp+ SexOfChild+ age_sib+ years_at_risk, data = h
ealthySib, sum)
  healthySib_aggr<- subset(healthySib_aggr, SexOfChild!=9)
  healthySib_aggr$Sex<- recode_factor(healthySib_aggr$SexOfChild, "1" = "Male", "2" = "F
emale")
  healthySib_aggr$age_start_risk<- healthySib_aggr$age_sib- healthySib_aggr$years_at_ris
k
  healthySib_aggr<- subset(healthySib_aggr, years_at_risk>0)

  all(healthySib_aggr$age_sib>=healthySib_aggr$years_at_risk)

#############################################
# A data set with only proband and mother
  probands$status_new = "Proband"
```

```
  mothers$status_new = "Mother"
  PM<- rbind(probands, mothers)
  # year at risk of a mother = 2015- year of proband's diagnosis
  setDT(PM)[, years_at_risk := 2015- YEARDX[status_new=="Proband"], by = mother_id_num]
  # Age for healthy mothers= 2015- (kids YOB- Age of mother)
  setDT(PM)[, age_of_mother2015 := 2015- (birth_year[status_new=="Proband"]- AgeOfMother
[status_new=="Proband"]), by = mother_id_num]
  # Age start risk for a mother= mother's age at proband's diagnosis
   setDT(PM)[, age_start_risk_mother := YEARDX[status_new=="Proband"]- (birth_year[statu
s_new=="Proband"]- AgeOfMother[status_new=="Proband"]), by = mother_id_num]

  mothers = subset(PM, status_new== "Mother")
  mothers<- subset(mothers, age_of_mother2015<=26)
   # Recode age
  mothers$agegp[mothers$age_of_mother2015>=15&mothers$age_of_mother2015<=19]<- "15-19 ye
ars"
  mothers$agegp[mothers$age_of_mother2015>=20&mothers$age_of_mother2015<=24]<- "20-24 ye
ars"
  mothers$agegp[mothers$age_of_mother2015>=25&mothers$age_of_mother2015<=29]<- "25-29 ye
ars"
  mothers= mothers%>%
  mutate(agegp_num= ifelse(agegp=="15-19 years", 1, ifelse(agegp=="20-24 years", 2, ifel
se(agegp=="25-29 years", 3, NA))))

  # Include only healthy mother to the denominator
  summary(mothers$PATIENT_ID_num)
  healthyMothers<- mothers[is.na(mothers$PATIENT_ID_num), ]

  ## find the age distribution at yeardx for affected mothers
  motherCasesdf1<- mothers[!is.na(mothers$PATIENT_ID_num), ]
  # motherCasesdf1<- subset(motherCasesdf1, AGE>=14)
  motherCasesdf1<- motherCasesdf1[order(motherCasesdf1$YEARDX), ]
  motherCasesdf1<- distinct(motherCasesdf1, PATIENT_ID_num, .keep_all= TRUE) #Keep the f
irst record for each mother

  motherCasesdf1= motherCasesdf1%>%
  mutate(agegpDX= ifelse(AGE>=15&AGE<=19, "15-19 years", ifelse(AGE>=20&AGE<=24, "20-24
 years", ifelse(AGE>=25&AGE<=29, "25-29 years",  NA))))

  motherCasesdf1= motherCasesdf1%>%
  mutate(agegp_num= ifelse(agegpDX=="15-19 years", 1, ifelse(agegpDX=="20-24 years", 2,
 ifelse(agegpDX=="25-29 years", 3, NA))))

  motherCasesdf2<- subset(motherCasesdf1, agegpDX %in% c(healthyMothers$agegp))
  healthyMothers<- subset(healthyMothers, agegp %in% c(motherCasesdf2$agegpDX))

  # If there are healthy mothers, include them to the denominator
  if(nrow(healthyMothers) > 0){
    healthyMothers$Count<- 1
    healthyMothers_aggr<- aggregate(Count~ agegp+ years_at_risk+ age_start_risk_mother+
 age_of_mother2015, data = healthyMothers, sum)
    healthyMothers_aggr$sex<- "Female"
    healthyMothers_aggr<- subset(healthyMothers_aggr, years_at_risk>0)
    names(healthyMothers_aggr)<- c("agegp", "years_at_risk", "age_start_risk", "age2015"
```

```
, "Count", "sex")
    healthyMothers_aggr$status<- "Healthy mother"
  } else {
    healthyMothers_aggr<- data.frame(agegp = character(), years_at_risk = numeric(), age
_start_risk = numeric(), age2015 = numeric(), Count = numeric(), sex = character(), stat
us = character())
  }

  res<- data.frame(c("Proband", "Sibling", "Mother", "Mother and Sibling"),
  c(probandsN, sibCase_num_people, motherCases_num_people, sibCase_num_people+motherCase
s_num_people))
  names(res)<- c("No_of_people_with_cancer", "N")
  print(res)
  #############################################
  # merge healthy siblings and mothers
  healthySib_aggr<- subset(healthySib_aggr, select = c("agegp", "age_sib", "years_at_ris
k", "Count", "Sex", "age_start_risk"))
  names(healthySib_aggr)<- c("agegp", "age2015", "years_at_risk", "Count", "sex", "age_s
tart_risk")
  healthySib_aggr$status<- "Healthy sibling"
  healthy_mother_sib<- rbind(healthySib_aggr, healthyMothers_aggr)

write.csv(healthy_mother_sib, paste0(workdir, "df1.csv"), row.names = F)

}
```

```
# Everyone
all_races = unique(c(fambirthccrT$RaceEthnicityOfMother))
all_ethnicities = unique(c(fambirthccrT$HispanicOriginCodeOfMother))

any_cancer(icd$icd_codes, all_races, all_ethnicities)
```

```
##   No_of_people_with_cancer     N
## 1                  Proband 29072
## 2                  Sibling   112
## 3                   Mother    65
## 4       Mother and Sibling   177
```

```
# By cancer groups
# any_cancer(icd_list[[1]], all_races, all_ethnicities)
# Repeat for group 2 to 12/ hema/ solid
```

- Calculate the expected number of cases
  - Calculate person-years at risk
  - Age-adjusted incidence rate per 100,000 of all cancers ($\lambda_k$) derived from SEER 13 Regs Research Data, Nov 2018 Sub (1992-2016), California

```
import csv

def fill_weight(age_map, value, start, end):
    for i in range(start, end):
        age_map[i] = value

with open('df1.csv') as df1:
    df1_reader = csv.reader(df1, delimiter=',')
    df1_reader_list = list(df1_reader)
    #init weight map for age 0 ~ 99
    male_age_map = [0] * 100
    female_age_map = [0] * 100

    #fill weights into male_age_map here
    fill_weight(male_age_map, 26.01, 0, 1)
    fill_weight(male_age_map, 23.07, 1, 5)
    fill_weight(male_age_map, 13.3, 5, 10)
    fill_weight(male_age_map, 14.67, 10, 15)
    fill_weight(male_age_map, 24.05, 15, 20)
    fill_weight(male_age_map, 36.11, 20, 25)
    fill_weight(male_age_map, 53.7, 25, 30)

    #fill weights into female_age_map here
    fill_weight(female_age_map, 25.27, 0, 1)
    fill_weight(female_age_map, 20.04, 1, 5)
    fill_weight(female_age_map, 11.4, 5, 10)
    fill_weight(female_age_map, 13.74, 10, 15)
    fill_weight(female_age_map, 23.24, 15, 20)
    fill_weight(female_age_map, 39.2, 20, 25)
    fill_weight(female_age_map, 63.13, 25, 30)
    fill_weight(female_age_map, 106.16, 30, 35)
    fill_weight(female_age_map, 172.88, 35, 40)
    fill_weight(female_age_map, 300.01, 40, 45)
    fill_weight(female_age_map, 451.87, 45, 50)
    fill_weight(female_age_map, 605.23, 50, 55)
    fill_weight(female_age_map, 771.12, 55, 60)
    fill_weight(female_age_map, 987.88, 60, 65)
    fill_weight(female_age_map, 1248.26, 65, 70)
    fill_weight(female_age_map, 1466.16, 70, 75)
    fill_weight(female_age_map, 1653.46, 75, 80)
    fill_weight(female_age_map, 1763.71, 80, 85)

    # print(male_age_map, female_age_map)

    hash_map = {"Male": male_age_map, "Female": female_age_map}
    for i, row in enumerate(df1_reader_list):
        if i == 0:
            df1_reader_list[i].append("result")
        else:
            current_age = int(row[1])
            count = int(row[3])
            sex = row[4]
            start_age = int(row[5])
```

```
            result = 0
            for j in range(start_age, current_age + 1):
                if j == start_age or j == current_age:
                    result += 0.5 * hash_map[sex][j]
                else:
                    result += hash_map[sex][j]
            df1_reader_list[i].append(round(count * result, 2))
    with open('result.csv', 'w') as f:
        writer = csv.writer(f, delimiter=',')
        writer.writerows(df1_reader_list)
```

Reference for confidence interval calculation:
https://www.doh.wa.gov/Portals/1/Documents/1500/ConfIntGuide.pdf
(https://www.doh.wa.gov/Portals/1/Documents/1500/ConfIntGuide.pdf)

```
# Calculate SIR and CI
## observed
O = 177 # O = No. of sibling cases+ No. of mother cases
## expected
result<- read.csv(paste0(workdir, "result.csv"), sep = ",", header = T)
E= sum(result$result)/100000
## SIR
sir1= O/E

# Confidence interval
## For observed <= 100 cases, use Poisson approximation
# poisson_table<- read.delim(paste0(workdir, "poisson_table.txt"), sep = "", header = T)
#
# poisson_table1<- subset(poisson_table, Observed == O)
# LL_small= poisson_table1$Lower_limit/E
# UL_small= poisson_table1$Upper_limit/E

## For observed >100 cases, use the equation suggested by Washington State Department of
Health
LL_large = (1- (1/(9*O))- (1.96/(3*sqrt(O))))^3 * (O/E)
UL_large = (1- (1/(9*(O+1)))+ (1.96/(3*sqrt(O+1))))^3 * ((O+1)/E)

dfr<- data.frame(O, E, paste0(round(sir1,2), " (", round(LL_large, 2), ", ", round(UL_la
rge, 2), ")"))
names(dfr)<- c("Observed", "Expected", "SIR (95%CI)")
dfr
```

```
##   Observed Expected       SIR (95%CI)
## 1      177 53.24353 3.32 (2.85, 3.85)
```

### 1.2 Calculate the SIR of affected mother/sibling of any cancer by race/ethnic group

#### Non-Latino White

```
NLW_race = 10
NLW_ethnicity = 1
# Any cancer
any_cancer(icd$icd_codes, NLW_race, NLW_ethnicity)
```

```
##   No_of_people_with_cancer    N
## 1                  Proband 8119
## 2                  Sibling   33
## 3                   Mother   17
## 4       Mother and Sibling   50
```

```
# By cancer groups
# any_cancer(icd_list[[1]], NLW_race, NLW_ethnicity)
# Repeat for group 2 to 12/ hema/ solid
```

- Calculate the expected number of cases
    - Calculate person-years at risk
    - Age-adjusted incidence rate per 100,000 of all cancers ($\lambda_k$) derived from SEER 13 Regs Research Data, Nov 2018 Sub (1992-2016), California, Non-Hispanic/Latino White subjects only

```
import csv

def fill_weight(age_map, value, start, end):
    for i in range(start, end):
        age_map[i] = value

with open('df1.csv') as df1:
    df1_reader = csv.reader(df1, delimiter=',')
    df1_reader_list = list(df1_reader)
    #init weight map for age 0 ~ 99
    male_age_map = [0] * 100
    female_age_map = [0] * 100

    #fill weights into male_age_map here
    fill_weight(male_age_map, 29.59, 0, 1)
    fill_weight(male_age_map, 24.26, 1, 5)
    fill_weight(male_age_map, 14.97, 5, 10)
    fill_weight(male_age_map, 15.79, 10, 15)
    fill_weight(male_age_map, 28.3, 15, 20)
    fill_weight(male_age_map, 46.56, 20, 25)
    fill_weight(male_age_map, 67.74, 25, 30)

    #fill weights into female_age_map here
    fill_weight(female_age_map, 23.06, 0, 1)
    fill_weight(female_age_map, 21, 1, 5)
    fill_weight(female_age_map, 13.1, 5, 10)
    fill_weight(female_age_map, 13.7, 10, 15)
    fill_weight(female_age_map, 28.6, 15, 20)
    fill_weight(female_age_map, 52.73, 20, 25)
    fill_weight(female_age_map, 79.89, 25, 30)


    # print(male_age_map, female_age_map)

    hash_map = {"Male": male_age_map, "Female": female_age_map}
    for i, row in enumerate(df1_reader_list):
        if i == 0:
            df1_reader_list[i].append("result")
        else:
            current_age = int(row[1])
            count = int(row[3])
            sex = row[4]
            start_age = int(row[5])
            result = 0
            for j in range(start_age, current_age + 1):
                if j == start_age or j == current_age:
                    result += 0.5 * hash_map[sex][j]
                else:
                    result += hash_map[sex][j]
            df1_reader_list[i].append(round(count * result, 2))
    with open('result.csv', 'w') as f:
        writer = csv.writer(f, delimiter=',')
        writer.writerows(df1_reader_list)
```

Reference for confidence interval calculation:
https://www.doh.wa.gov/Portals/1/Documents/1500/ConfIntGuide.pdf
(https://www.doh.wa.gov/Portals/1/Documents/1500/ConfIntGuide.pdf)

```
# Calculate SIR and CI
## observed
O = 50 # O = No. of sibling cases+ No. of mother cases
## expected
result<- read.csv(paste0(workdir, "result.csv"), sep = ",", header = T)
E= sum(result$result)/100000
## SIR
sir1= O/E

# Confidence interval
## For observed <= 100 cases, use Poisson approximation
poisson_table<- read.delim(paste0(workdir, "poisson_table.txt"), sep = "", header = T)

poisson_table1<- subset(poisson_table, Observed == O)
LL_small= poisson_table1$Lower_limit/E
UL_small= poisson_table1$Upper_limit/E

## For observed >100 cases, use the equation suggested by Washington State Department of
Health
# LL_large = (1- (1/(9*O))- (1.96/(3*sqrt(O))))^3 * (O/E)
# UL_large = (1- (1/(9*(O+1)))+ (1.96/(3*sqrt(O+1))))^3 * ((O+1)/E)

dfr<- data.frame(O, E, paste0(round(sir1,2), " (", round(LL_small, 2), ", ", round(UL_sm
all, 2), ")"))
names(dfr)<- c("Observed", "Expected", "SIR (95%CI)")
dfr
```

```
##   Observed Expected      SIR (95%CI)
## 1       50 19.24007 2.6 (1.93, 3.43)
```

### Latino all races

```
all_races = unique(c(fambirthccrT$RaceEthnicityOfMother))
Latino_ethnicity = c(2,3,4,5,6,8)
# Any cancer
any_cancer(icd$icd_codes, all_races, Latino_ethnicity)
```

```
##   No_of_people_with_cancer     N
## 1                  Proband 12736
## 2                  Sibling    52
## 3                   Mother    26
## 4       Mother and Sibling    78
```

```
# By cancer groups
# any_cancer(icd_list[[1]], all_races, Latino_ethnicity)
# Repeat for group 2 to 12/ hema/ solid
```

- Calculate the expected number of cases
    - Calculate person-years at risk
    - Age-adjusted incidence rate per 100,000 of all cancers ($\lambda_k$) derived from SEER 13 Regs Research Data, Nov 2018 Sub (1992-2016), California, Hispanic/Latino subjects of all races only

```
import csv

def fill_weight(age_map, value, start, end):
    for i in range(start, end):
        age_map[i] = value

with open('df1.csv') as df1:
    df1_reader = csv.reader(df1, delimiter=',')
    df1_reader_list = list(df1_reader)
    #init weight map for age 0 ~ 99
    male_age_map = [0] * 100
    female_age_map = [0] * 100

    #fill weights into male_age_map here
    fill_weight(male_age_map, 24.08, 0, 1)
    fill_weight(male_age_map, 22.99, 1, 5)
    fill_weight(male_age_map, 12.95, 5, 10)
    fill_weight(male_age_map, 14.82, 10, 15)
    fill_weight(male_age_map, 23.86, 15, 20)
    fill_weight(male_age_map, 31.28, 20, 25)
    fill_weight(male_age_map, 40.16, 25, 30)

    #fill weights into female_age_map here
    fill_weight(female_age_map, 22.63, 0, 1)
    fill_weight(female_age_map, 20.31, 1, 5)
    fill_weight(female_age_map, 11.86, 5, 10)
    fill_weight(female_age_map, 13.77, 10, 15)

    fill_weight(female_age_map, 20.39, 15, 20)
    fill_weight(female_age_map, 33.08, 20, 25)
    fill_weight(female_age_map, 53.77, 25, 30)
    fill_weight(female_age_map, 91.05, 30, 35)
    fill_weight(female_age_map, 143.25, 35, 40)

    # print(male_age_map, female_age_map)

    hash_map = {"Male": male_age_map, "Female": female_age_map}
    for i, row in enumerate(df1_reader_list):
        if i == 0:
            df1_reader_list[i].append("result")
        else:
            current_age = int(row[1])
            count = int(row[3])
            sex = row[4]
            start_age = int(row[5])
            result = 0
            for j in range(start_age, current_age + 1):
                if j == start_age or j == current_age:
                    result += 0.5 * hash_map[sex][j]
                else:
                    result += hash_map[sex][j]
            df1_reader_list[i].append(round(count * result, 2))
    with open('result.csv', 'w') as f:
```

```
        writer = csv.writer(f, delimiter=',')
        writer.writerows(df1_reader_list)
```

Reference for confidence interval calculation:
https://www.doh.wa.gov/Portals/1/Documents/1500/ConfIntGuide.pdf
(https://www.doh.wa.gov/Portals/1/Documents/1500/ConfIntGuide.pdf)

```
# Calculate SIR and CI
## observed
O = 78 # O = No. of sibling cases+ No. of mother cases
## expected
result<- read.csv(paste0(workdir, "result.csv"), sep = ",", header = T)
E= sum(result$result)/100000
## SIR
sir1= O/E

# Confidence interval
## For observed <= 100 cases, use Poisson approximation
poisson_table<- read.delim(paste0(workdir, "poisson_table.txt"), sep = "", header = T)

poisson_table1<- subset(poisson_table, Observed == O)
LL_small= poisson_table1$Lower_limit/E
UL_small= poisson_table1$Upper_limit/E

## For observed >100 cases, use the equation suggested by Washington State Department of
Health
# LL_large = (1- (1/(9*O))- (1.96/(3*sqrt(O))))^3 * (O/E)
# UL_large = (1- (1/(9*(O+1)))+ (1.96/(3*sqrt(O+1))))^3 * ((O+1)/E)

dfr<- data.frame(O, E, paste0(round(sir1,2), " (", round(LL_small, 2), ", ", round(UL_sm
all, 2), ")"))
names(dfr)<- c("Observed", "Expected", "SIR (95%CI)")
dfr
```

```
##   Observed Expected       SIR (95%CI)
## 1       78 23.21456 3.36 (2.66, 4.19)
```

### 1.3 Compare the SIR of affected mother/sibling of any cancer by race/ethnic group with an approximate Chi-square test

We compared the SIRs across race/ethnic groups with approximate Chi-squared tests. The approximate chi-square method compares the probability of occurrence of events in one group to another, based on a binomial distribution. This comparison is not related to the 95% confidence intervals for the SIRs which are based on a Poisson distribution and may overlap between two groups that are significantly different by approximate chi-square comparison.

Below is a detailed demonstration. For example, for the overall category, the SIR = 2.60 (95%CI: 1.66, 4.06) (Observed No. of affected relatives = 50; Expected No. of affected relatives = 19.24) for non-Latino White subjects and SIR = 3.35 (95%CI: 2.24, 5.05) (Observed No. of affected relatives = 78; Expected No. of affected relatives = 23.22) for Latino subjects.

With the following approximation method:

#### Approximate test for $H_0$: $\psi=1$:

$$\frac{(D_2 - E(D_2))^2}{Var(D_2)} \sim \chi_1^2$$

To get an expression for $E(D_2)$ and $Var(D_2)$, we use the binomial distribution of $D_2$ (conditioning on the total number of events, $D_+$) as before.

In a binomial distribution, $E(D_2)$ = total events * probability of event

$$= D_+\pi = D_+\left(\frac{E_2}{E_+}\right) = \tilde{E}_2 \quad \text{(recall that under } H_0,\ \pi = \frac{E_2}{E_+}\text{)}$$

and $Var(D_2)$ $= D_+\pi(1-\pi) = D_+\left(\frac{E_2}{E_+}\right)\left(1-\frac{E_2}{E_+}\right) = \tilde{E}_2\left(\frac{E_1}{E_+}\right)$

$$\tilde{E}_2\left(\frac{D_+}{D_+}\right)\left(\frac{E_1}{E_+}\right) = \frac{\tilde{E}_2\tilde{E}_1}{D_+} = \frac{\tilde{E}_1\tilde{E}_2}{\tilde{E}_1 + \tilde{E}_2}$$

Substituting in these expressions for $E(D_2)$ and $Var(D_2)$ in the formula for the approximate test above, we can rewrite the approximate test as:

$$\frac{(D_2 - \tilde{E}_2)^2(\tilde{E}_1 + \tilde{E}_2)}{\tilde{E}_1\tilde{E}_2} = \frac{\tilde{E}_1(D_2 - \tilde{E}_2)^2 + \tilde{E}_2(D_2 - \tilde{E}_2)^2}{\tilde{E}_1\tilde{E}_2}$$

(NOTE: Since $\begin{array}{l} D_+ = \tilde{E}_+, \\ D_1 + D_2 = \tilde{E}_1 + \tilde{E}_2, \\ D_2 - \tilde{E}_2 = -(D_1 - \tilde{E}_1)) \end{array}$

So, $$\frac{\widetilde{E}_1\left(D_2-\widetilde{E}_2\right)^2+\widetilde{E}_2\left(D_2-\widetilde{E}_2\right)^2}{\widetilde{E}_1\widetilde{E}_2}=\frac{\widetilde{E}_1\left(D_2-\widetilde{E}_2\right)^2+\widetilde{E}_2\left(D_1-\widetilde{E}_1\right)^2}{\widetilde{E}_1\widetilde{E}_2}$$

$$=\frac{\left(D_1-\widetilde{E}_1\right)^2}{\widetilde{E}_1}+\frac{\left(D_2-\widetilde{E}_2\right)^2}{\widetilde{E}_2}\sim\chi_1^2$$

This is the approximate test for $H_0$: $\psi=1$

```
comp_chi2<- function(d2, e2_star, d1, e1_star){
  d       = d2+d1
  e_star  = e2_star+e1_star
  e2_star_tilde = d*e2_star/e_star
  e1_star_tilde = d*e1_star/e_star
# -- chisq test based on eq 3.7 in B&D;
  chisq = (abs(d1 - e1_star_tilde) - 0.5)^2 / e1_star_tilde +
    (abs(d2 - e2_star_tilde) - 0.5)^2 / e2_star_tilde
                                                # p = 1 - probchi(chisq,1)
  p = pchisq(chisq, df=1, lower.tail=FALSE)
  print(p)
}

  # -- group 2 is  non-Latino White;
  d2      = 50
  e2_star = 19.24007

  # -- group 1 is  Latino;
  d1      =  78
  e1_star =  23.21456

paste0("p-value comparing the SIR of NLW to Latino: ")
```

```
## [1] "p-value comparing the SIR of NLW to Latino: "
```

```
comp_chi2(d2, e2_star, d1, e1_star)
```

```
## [1] 0.1824724
```

### 2. SIR of the same cancer with the proband

The SIR for affected relatives with the same group of cancer with the proband was calculated in a similar manner with the SIR of any cancer.

```
same_cancer <- function(icd, race, ethnicity){
  cases<- subset(fambirthccrTcases, HISTO_T3 %in% icd & RaceEthnicityOfMother %in% race
 & HispanicOriginCodeOfMother %in% ethnicity)

  # Restrict yeardx to 1989-2015
  cases<- subset(cases, YEARDX>=1989& YEARDX<=2015)
  casesN<- nrow(cases) # Count number of cases

  # Find the probands
  ## Probands are any kid who got the case, one family can only have one proband (case o
f the earliest yeardx)
  probands_pre<- completeFun(cases, c("vitstat_childID_num")) # Extract child cases
  probands_pre<- probands_pre[order(probands_pre$YEARDX), ] # Sort by yeardx
  probands<- distinct(probands_pre, mother_id_num, .keep_all= TRUE) # Keep only one chil
d for each family
  fams<- subset(fambirthccrT, mother_id_num %in% c(probands$mother_id_num))
  probandsN<- nrow(data.frame(table(probands$vitstat_childID_num))) # count number of pr
obands

#############################################
  # Count the number of mother cases of any cancer, diagnosed under 26
  mothers<- subset(fams, status == "Mother")
  motherCasesdf<- completeFun(mothers, c("PATIENT_ID_num"))
  # Find mother cases of the same group
  motherCasesdf<- subset(motherCasesdf, HISTO_T3 %in% icd & AGE<=26)
  motherCases_num_rec<- nrow(motherCasesdf)
  motherCases_num_people<- nrow(data.frame(table(motherCasesdf$PATIENT_ID_num)))
  ############ motherCases_num_people is part of the numerator ############

#############################################
  # Count the number of sibling cases of any cancer
  childCases<- completeFun(fams, c("vitstat_childID_num", "PATIENT_ID_num"))
  sibCasesdf<- subset(childCases, !(childCases$PATIENT_ID_num %in% c(probands$PATIENT_ID
_num)))
  # Find sibling cases of the same group
  sibCasesdf<- subset(sibCasesdf, HISTO_T3 %in% icd)
  sibCase_num_rec<- nrow(sibCasesdf)
  sibCase_num_people<- nrow(data.frame(table(sibCasesdf$PATIENT_ID_num)))
  ############ sibCase_num_people is part of the numerator ############

#############################################
#Find healthy siblings to calculate the denominator
  children<- completeFun(fams, c("vitstat_childID_num"))
  allSib<- subset(children, !(PATIENT_ID_num %in% c(probands$PATIENT_ID_num)))
  #all Siblings (including healthy)

# year at risk of a sibling=
## 1) 2015- year of proband's diagnosis if sibling was born before proband was diagnosed
## 2) 2015- birth_year of sibling if sibling was born after proband was diagnosed
  probands_1<- probands[order(probands$mother_id, probands$AGE), ]
  sum(unique(length(probands_1$mother_id_num)))
  probands_1<- probands_1[!duplicated(probands_1$mother_id_num),]
  probands_1$status_new<- "Proband"
```

```
  allSib$status_new<- "Sibling"

  children_1<- rbind(probands_1, allSib)
  children_1<- children_1[order(children_1$mother_id_num), ]

  setDT(children_1)[, risk_status := birth_year- YEARDX[status_new=="Proband"], by = mot
her_id_num]
  children_1$risk_statusN[children_1$risk_status<=0]<-1 #birth_year<=yeardx
  children_1$risk_statusN[children_1$risk_status>0]<-2 #birth_year>yeardx

  #1)
  children_11<- subset(children_1, risk_statusN==1)
  setDT(children_11)[, years_at_risk := 2015- YEARDX[status_new=="Proband"], by = mother
_id_num]
  setDT(children_11)[, AgeDiff := birth_year- birth_year[status_new=="Proband"], by = mo
ther_id_num]
  children_11Sib<- subset(children_11, status_new=="Sibling")
#2)
  children_12<- subset(children_1, risk_statusN==2)
  children_12$years_at_risk<- 2015-children_12$birth_year
  children_12$AgeDiff<- 0
  children_1<- rbind(children_11, children_12)

  # Crop out siblings
  allSib<- subset(children_1, status_new=="Sibling")
  allSib$age_sib<- 2015-allSib$birth_year

  # Recode age
  allSib$agegp[allSib$age_sib==0]<- "00 years"
  allSib$agegp[allSib$age_sib>=1&allSib$age_sib<=4]<- "01-04 years"
  allSib$agegp[allSib$age_sib>=5&allSib$age_sib<=9]<- "05-09 years"
  allSib$agegp[allSib$age_sib>=10&allSib$age_sib<=14]<- "10-14 years"
  allSib$agegp[allSib$age_sib>=15&allSib$age_sib<=19]<- "15-19 years"
  allSib$agegp[allSib$age_sib>=20&allSib$age_sib<=24]<- "20-24 years"
  allSib$agegp[allSib$age_sib>=25]<- "25-29 years"

  # Keep only healthy siblings to the denominator
  summary(allSib$PATIENT_ID)
  healthySib<- subset(allSib, !(vitstat_childID_num %in% c(sibCasesdf$vitstat_childID_nu
m)))
  healthySib$Count<- 1
  healthySib_aggr<- aggregate(Count~ agegp+ SexOfChild+ age_sib+ years_at_risk, data = h
ealthySib, sum)
  healthySib_aggr<- subset(healthySib_aggr, SexOfChild!=9)
  healthySib_aggr$Sex<- recode_factor(healthySib_aggr$SexOfChild, "1" = "Male", "2" = "F
emale")
  healthySib_aggr$age_start_risk<- healthySib_aggr$age_sib- healthySib_aggr$years_at_ris
k
  healthySib_aggr<- subset(healthySib_aggr, years_at_risk>0)

  all(healthySib_aggr$age_sib>=healthySib_aggr$years_at_risk)

##############################################
# A data set with only proband and mother
```

```
  probands$status_new = "Proband"
  mothers$status_new = "Mother"
  PM<- rbind(probands, mothers)
  # year at risk of a mother = 2015- year of proband's diagnosis
  setDT(PM)[, years_at_risk := 2015- YEARDX[status_new=="Proband"], by = mother_id_num]
  # Age for healthy mothers= 2015- (kids YOB- Age of mother)
  setDT(PM)[, age_of_mother2015 := 2015- (birth_year[status_new=="Proband"]- AgeOfMother
[status_new=="Proband"]), by = mother_id_num]
  # Age start risk for a mother= mother's age at proband's diagnosis
   setDT(PM)[, age_start_risk_mother := YEARDX[status_new=="Proband"]- (birth_year[statu
s_new=="Proband"]- AgeOfMother[status_new=="Proband"]), by = mother_id_num]

  mothers = subset(PM, status_new== "Mother")
  mothers<- subset(mothers, age_of_mother2015<=26)
   # Recode age
  mothers$agegp[mothers$age_of_mother2015>=15&mothers$age_of_mother2015<=19]<- "15-19 ye
ars"
  mothers$agegp[mothers$age_of_mother2015>=20&mothers$age_of_mother2015<=24]<- "20-24 ye
ars"
  mothers$agegp[mothers$age_of_mother2015>=25&mothers$age_of_mother2015<=29]<- "25-29 ye
ars"
  mothers= mothers%>%
  mutate(agegp_num= ifelse(agegp=="15-19 years", 1, ifelse(agegp=="20-24 years", 2, ifel
se(agegp=="25-29 years", 3, NA))))

  # Include only healthy mother to the denominator
  summary(mothers$PATIENT_ID_num)
  healthyMothers<- mothers[is.na(mothers$PATIENT_ID_num), ]

  ## find the age distribution at yeardx for affected mothers
  motherCasesdf1<- mothers[!is.na(mothers$PATIENT_ID_num), ]
  # motherCasesdf1<- subset(motherCasesdf1, AGE>=14)
  motherCasesdf1<- motherCasesdf1[order(motherCasesdf1$YEARDX), ]
  motherCasesdf1<- distinct(motherCasesdf1, PATIENT_ID_num, .keep_all= TRUE) #Keep the f
irst record for each mother

  motherCasesdf1= motherCasesdf1%>%
  mutate(agegpDX= ifelse(AGE>=15&AGE<=19, "15-19 years", ifelse(AGE>=20&AGE<=24, "20-24
 years", ifelse(AGE>=25&AGE<=29, "25-29 years",  NA))))

  motherCasesdf1= motherCasesdf1%>%
  mutate(agegp_num= ifelse(agegpDX=="15-19 years", 1, ifelse(agegpDX=="20-24 years", 2,
 ifelse(agegpDX=="25-29 years", 3, NA))))

  motherCasesdf2<- subset(motherCasesdf1, agegpDX %in% c(healthyMothers$agegp))
  healthyMothers<- subset(healthyMothers, agegp %in% c(motherCasesdf2$agegpDX))

  # If there are healthy mothers, include them to the denominator
  if(nrow(healthyMothers) > 0){
    healthyMothers$Count<- 1
    healthyMothers_aggr<- aggregate(Count~ agegp+ years_at_risk+ age_start_risk_mother+
 age_of_mother2015, data = healthyMothers, sum)
    healthyMothers_aggr$sex<- "Female"
    healthyMothers_aggr<- subset(healthyMothers_aggr, years_at_risk>0)
```

```
    names(healthyMothers_aggr)<- c("agegp", "years_at_risk", "age_start_risk", "age2015"
, "Count", "sex")
    healthyMothers_aggr$status<- "Healthy mother"
  } else {
    healthyMothers_aggr<- data.frame(agegp = character(), years_at_risk = numeric(), age
_start_risk = numeric(), age2015 = numeric(), Count = numeric(), sex = character(), stat
us = character())
  }

  res<- data.frame(c("Proband", "Sibling", "Mother", "Mother and Sibling"),
  c(probandsN, sibCase_num_people, motherCases_num_people, sibCase_num_people+motherCase
s_num_people))
  names(res)<- c("No_of_people_with_cancer", "N")
  print(res)
  #############################################
  # merge healthy siblings and mothers
  healthySib_aggr<- subset(healthySib_aggr, select = c("agegp", "age_sib", "years_at_ris
k", "Count", "Sex", "age_start_risk"))
  names(healthySib_aggr)<- c("agegp", "age2015", "years_at_risk", "Count", "sex", "age_s
tart_risk")
  healthySib_aggr$status<- "Healthy sibling"
  healthy_mother_sib<- rbind(healthySib_aggr, healthyMothers_aggr)

write.csv(healthy_mother_sib, paste0(workdir, "df2.csv"), row.names = F)

}
```

```
# Everyone
all_races = unique(c(fambirthccrT$RaceEthnicityOfMother))
all_ethnicities = unique(c(fambirthccrT$HispanicOriginCodeOfMother))

same_cancer(icd_list[[1]], all_races, all_ethnicities)
```

```
##   No_of_people_with_cancer    N
## 1                  Proband 8500
## 2                  Sibling    8
## 3                   Mother    3
## 4       Mother and Sibling   11
```

```
# Repeat for group 2 to 12/ hema/ solid
```

Person-years of Group 1 cancer (Leukemias)

- Calculate the expected number of cases
  - Calculate person-years at risk
  - Age-adjusted incidence rate per 100,000 of leukemias ($\lambda_k$, ICCC group 1) derived from SEER 13 Regs Research Data, Nov 2018 Sub (1992-2016), California.

```
import csv
with open('df2.csv') as df2:
    df2_reader = csv.reader(df2, delimiter=',')
    df2_reader_list = list(df2_reader)
    male = [6.02] + [11.03] * 4 + [5.3] * 5 + [4.35] * 5 + [4.55] * 5 + [3.34] * 5 + [3.
25] * 5
    female = [4.99] + [9.06] * 4 + [4.55] * 5 + [3.1] * 5 + [3.05] * 5 + [2.52] * 5 + [
2.78] * 5
    hash_map = {"Male": male, "Female": female}
    for i, row in enumerate(df2_reader_list):
        if i == 0:
            df2_reader_list[i].append("result2")
        else:
            current_age = int(row[1])
            count = int(row[3])
            sex = row[4]
            start_age = int(row[5])
            result2 = 0
            for j in range(start_age, current_age + 1):
                if j == start_age or j == current_age:
                    result2 += 0.5 * hash_map[sex][j]
                else:
                    result2 += hash_map[sex][j]
            df2_reader_list[i].append(round(count * result2, 2))
    with open('result2.csv', 'w') as f:
        writer = csv.writer(f, delimiter=',')
        writer.writerows(df2_reader_list)
```

```
# Calculate SIR and CI
## observed
O = 11 # O = No. of sibling cases+ No. of mother cases
## expected
result2<- read.csv(paste0(workdir, "result2.csv"), sep = ",", header = T)
E= sum(result2$result2)/100000
## SIR
sir1= O/E

# Confidence interval
## For observed <= 100 cases, use Poisson approximation
poisson_table<- read.delim(paste0(workdir, "poisson_table.txt"), sep = "", header = T)

poisson_table1<- subset(poisson_table, Observed == O)
LL_small= poisson_table1$Lower_limit/E
UL_small= poisson_table1$Upper_limit/E

## For observed >100 cases, use the equation suggested by Washington State Department of
Health
# LL_large = (1- (1/(9*O))- (1.96/(3*sqrt(O))))^3 * (O/E)
# UL_large = (1- (1/(9*(O+1)))+ (1.96/(3*sqrt(O+1))))^3 * ((O+1)/E)

dfr<- data.frame(O, E, paste0(round(sir1,2), " (", round(LL_small, 2), ", ", round(UL_sm
all, 2), ")"))
names(dfr)<- c("Observed", "Expected", "SIR (95%CI)")
dfr
```

```
##   Observed Expected       SIR (95%CI)
## 1       11 4.427232 2.48 (1.24, 4.45)
```

This process was repeated for group 2 to 12 of cancers, and hematologic cancers.

The SIRs by race/ethnic group were also calculated in the same manner.

### 3. SIR of second primary malignancies

```
rid<- read.csv("/Volumes/projects_wiemels/V_Foundation/Re-identification_12 broad groups
1010.csv", sep=",", header = T) # The file with random ID # Loaded to link subjects back

## Find second primaries
## Diagnosis of first primary: 1989-2015
df1<- read_xlsx("/Volumes/projects_wiemels/V_Foundation/SenkeiF/12 broad groups/Second p
rimaries/df1.xlsx") # The file processed by a physician (E. N.) that includes informatio
n on second primary malignancies
```

```
## New names:
## * status -> status...5
## * status -> status...9
```

```
df1$include<- as.numeric(as.character(df1$include))
df1$Dmother_id<- as.numeric(as.character(df1$Dmother_id))
df1$Dchild_id<- as.numeric(as.character(df1$Dchild_id))
df1$Dpatient_id<- as.numeric(as.character(df1$Dpatient_id))

# Second primaries: 1) case_cat= multiple records or intersection 2) include= 1 3) the k
id must have multiple records
df1<- subset(df1, case_cat!= "Sibling case")
df2<- completeFun(df1, c("include")) #ID of families included
df3<- subset(df1, Dmother_id %in% c(df2$Dmother_id)) # Families included

# Find child patients appeared more than once
df4<- completeFun(df3, c("Dpatient_id", "Dchild_id"))
dfx<- data.frame(table(df4$Dpatient_id))
dfx$Freq<- as.numeric(as.character(dfx$Freq))
dfx$Var1<- as.numeric(as.character(dfx$Var1))
dfx<- subset(dfx, Freq>= 2)
df4<- subset(df4, Dpatient_id %in% c(dfx$Var1))

# Find the first primary malignancy for each patient
df4<- df4[order(df4$Dpatient_id, df4$YEARDX), ] # order by yeardx
df5<- distinct(df4, Dpatient_id, .keep_all= TRUE) #Keep only one record for patient
df5$HISTO_T3<- as.numeric(as.character(df5$HISTO_T3))
```

```
second_primary<- function(icd){
  # Count number of second primaries in probands by icd group: secN
  df6<- subset(df5, HISTO_T3 %in% icd)
  secN<- nrow(data.frame(table(df6$Dpatient_id))) #Assuming each patient had only one se
cond primary
  ## Find original mother ids
  rid1<- subset(rid, Dmother_id %in% c(df6$Dmother_id))
  fams<- subset(fambirthccrT, mother_id_num %in% c(rid1$mother_id_num))
  probands<- subset(fams, PATIENT_ID_num %in% c(rid1$PATIENT_ID_num))

print(secN)
}
second_primary(icd$icd_codes)
```

```
## [1] 367
```

The second primary malignancies were reviewed by a physician (E.N.) to prevent the misclassification of relapsed first primary malignancies as second primary malignancies.

In addition, we identified 20 subjects with a third primary malignancy in total. So the total number of second primary malignancies was added up to 387.

The expected number of cases is the same as that calculated in # 1.1.
